# Dispelling stigmatizing misconceptions: A cross-sectional analysis of the impact of the adverse childhood experience of forced sexual intercourse on high-risk sexual behavior and weight status among adolescent girls

**DOI:** 10.1101/2022.11.08.22281983

**Authors:** Bethany Wrye, Poliala Dickson, Nickie M. D. Brillon, Angela S. Bowman

## Abstract

Although early sexual initiation (ESI) and elevated body mass index (BMI) are widely considered a threat to positive health, the misconception that elevated BMI is predictive of ESI produces additional stigmatizing burden for overweight or obese adolescent girls. This study expands on prior findings by exploring mediating impact of the adverse childhood experience (ACE) of forced sexual intercourse on BMI and ESI. Data from the U.S. population-based 2019 National Youth Risk Behavior Surveillance System (YRBS) were analyzed using complex samples analytic methods. Girls who reported forced intercourse experience and were Black or African American were more likely to report ESI, while those who were overweight or obese were no more likely to have engaged in ESI than healthy weight girls, when controlling for other ESI related risk-factors. Among girls who never experienced forced intercourse, current and prior substance use were predictive of ESI. Findings indicate that weight status does not impact ESI when accounting for forced sexual experiences, while early exposure to substance use may be a leverage point for intervention to reduce ESI among girls with prior experience of force. When developing interventions designed to reduce ESI, it is important to understand and account for the impact of ACEs.

## Introduction

Early sexual initiation (ESI) is widely considered to be a threat to positive health outcomes, particularly among adolescent girls (Dupere et al., 2008, Kaltiala-Heino et al., 2001). Early initiation of sex has been associated with transmission of sexually transmitted infections (Kaestle et al., 2005), teen pregnancy, and multiple sexual partners among others (Greenberg et al., 1992). Neighborhood poverty, conduct problems, and eating disorders have also been associated with ESI (Dupere et al., 2008, Kaltiala-Heino et al., 2001). While many factors have been associated with early sexual initiation, no consistent causal relationship has yet been established.

### Early sexual initiation as a high-risk sexual behavior

The definition of early initiation of sex or early sexual debut lacks consistency in literature. The most used definition is first sexual intercourse experience at age 14 or younger, others use the broad definition of sex at a young age or adolescence (Cavazos-Rehg et al., 2009; Kaplan et al., 2013). The Youth Risk Behavior Surveillance System (YRBSS) has a variable identifying participants who first had sexual intercourse before the age of 13 (CDC, 2018). The mean age of first sexual intercourse for girls in the United States is 17.3 years of age (National Center for Health Statistics, 2017). According to the CDC (2018), 40% of high school students have had sexual intercourse.

Price and Hyde (2001) found that several risk factors combine to have a cumulative predictive effect of ESI among adolescent girls. These include low self-esteem, poor parenting relationships, living in a non-intact household, higher levels of ADHD symptomology, low academic achievement, and low parental education level. Among young girls, early sexual experience was found to be associated with neighborhood poverty and history of conduct problems (Dupere et al., 2008), as well as bulimic-type eating pathology (Kaltiala-Heino et al., 2001). Substance use, particularly in the form of alcohol and marijuana, is a well-established risk factor for early sexual initiation (Little & Rankin, 2001). Girls are also more likely to report the initiation of sexual activity as a way of obtaining approval (Little & Rankin, 2001). Ellis et al. (2003) found that father absence is significantly associated with early sexual activity and adolescent pregnancy.

While many of the aforementioned factors, often described as delinquency, have been firmly established as being associated with early sexual initiation, a causal relationship is rarely discussed. Negriff et al. (2011) conducted a longitudinal study examining pubertal timing, sexual activity, and delinquency. Their findings indicated a temporal relationship where initiation of sexual activity precedes delinquency. It was also found that early pubertal timing, or early menarche for girls, precedes sexual activity. The authors posit that increased hormones associated with early pubertal timing led to an increased interest in sexual activity.

Early initiation of sex is associated with several health and social risks among young girls. Research indicates that early initiators are more likely to have multiple sexual partners, be involved in a teen pregnancy, teen birth, and having an abortion (Greenberg et al., 1992). Research has shown a relationship between early age of initiation of sex and sexually transmitted diseases. Kaestle et al.’s (2005) research indicates a strong relationship between young age of initiation of sex and odds of exposure to sexually transmitted infection. Previous research has indicated early sexual debut is one of the most robust predictors of sexually transmitted infection among adolescents and young adults (Greenberg et al., 1992).

### Adverse Childhood Experiences (ACEs)

The CDC-Kaiser ACE Study originally began in 1995. It includes questions measuring occurrence of ten experiences during the first 18 years of life: emotional abuse, physical abuse, sexual abuse, intimate partner violence, household problematic substance use, household mental illness, parental separation or divorce, and/or incarcerated household member. An estimated 65.5% of American women report having at least one ACE (CDC, 2016).

ACEs have been shown to have a dose dependent relationship with various health issues, including multiple sexual partners, sexually transmitted diseases, unintended pregnancies, early initiation of sexual activity, adolescent pregnancy, and risk for sexual violence (CDC, 2016). Kozak et al. (2018) found that adults who had experienced childhood sexual abuse were 1.7 times more likely to engage in delinquent and violent behavior. Cubellis et al. (2018) found similar results where childhood sexual abuse was significantly associated with perpetration of physical intimate partner violence.

Among girls who initiate sex before age 14, 70% have experienced forced intercourse (Greydanus et al., 2010). According to Hillis et al. (2001), as the number of ACEs experienced increased, the odds of a woman having reported their first sexual experience was prior to age 15 dramatically increased. They found that those who experienced one ACE were about twice as likely to have engaged in ESI, whereas those who experienced six or seven ACEs were as high as seven times more likely. McCutcheon et al. (2017) found parental alcohol use disorder and parental separation to be independent and significant predictors of early sexual debut, and early alcohol, tobacco, and cannabis use among offspring. Exposure to abuse or violence is also a risk factor for multiple sexual partners among adolescent girls. Silverman et al. (2011) found that adolescent girls who had experienced intimate partner violence were more likely to report multiple sexual partners, to report unprotected anal sex, partner sexual infidelity, fear of requesting condom use, negative consequences resulting from condom use requests, and failure to use condoms because of coercion. The health risks associated with multiple sexual partners coincide with most of those seen in early initiation of sex.

Recently, a study utilizing a large (n = 105,759) sample size of adolescents enrolled in public schools found ACEs and obesity to be strongly associated (Davis et al., 2019). The results of another study, “suggests that childhood maltreatment moderates the influence of genetic load for obesity on BMI as well as on the altered brain structure and function in reward related brain circuits” (Opel et al., 2019, p.18). In a study of adult women seeking weight loss therapy, Imperatori et al. (2016) found history of childhood trauma to be significantly associated with food addiction, binge eating, higher BMI, and anxiety/depressive symptoms. Isohookana et al. (2016) measured ACEs and BMI among adolescents aged 12-17 who had been admitted to an acute psychiatric hospital unit. They found that girls who had experienced sexual abuse were more likely to be obese.

### Body mass index (BMI)

As of 2014, 20.9% of American girls aged 12-19 are obese (Ogden et al., 2016). Among minority populations, the percentages are even higher, with 22.8% of Hispanic girls and 24.4% of Non-Hispanic Black or African American girls being categorized as obese. Potential reasons for the current high prevalence are varied and include health behaviors, the built environment, and socioeconomic status among many others (American College of Obstetricians and Gynecologists, 2017).

There are several health risks associated with obesity among adolescent girls, including impaired glucose tolerance (which may eventually lead to type 2 diabetes), cardiovascular disease in adulthood associated with hypertension and dyslipidemia, nonalcoholic fatty liver disease, disordered breathing, and orthopedic conditions (American College of Obstetricians and Gynecologists, 2017). Obesity also puts adolescent girls at increased risk for gynecological health issues related to increased levels of free estrogens and androgens, including abnormal uterine bleeding and polycystic ovary syndrome (American College of Obstetricians and Gynecologists, 2017).

Morbid obesity has been found to be associated with both early menarche (before 12 years of age) and teenage pregnancy (Neves et al., 2017). A meta-analysis of previous research examining this relationship revealed that early puberty was significantly more common among obese girls than girls of normal weight (Li et al., 2017). Negriff et al., (2011) found a mediational pathway such that early pubertal timing preceded sexual activity, which in turn preceded delinquency. These results indicate that early maturing adolescents may actively seek out sexual activity, leading to early sexual initiation. Neither friendship network characteristics nor maltreatment status were found to be significant. It may be possible that obesity results in early puberty, which leads to early sexual activity and eventually delinquency.

A study of adolescent girls seeking weight loss surgery due to severe obesity indicated that these girls had fewer sexual behaviors than their peers of a healthy weight (Becnel et al., 2016). Obesity may impact adolescent girls differently based on ethnicity. Leech and Dias (2012) found that for white adolescent girls, obesity was associated with multiple sex partners, failure to use condoms, and older sex partners. None of these factors were found to be significantly associated among African American girls.

While high risk sexual behaviors and BMI share several common risk factors, including ACEs. Previous studies examining the relationship between these factors could not be identified. This study seeks to examine the relationship between the ACE of forced sex, high risk sexual behaviors, and BMI among adolescent girls.

## Materials and Methods

The Youth Risk Behavior Surveillance System (YRBS) is an ongoing survey conducted by the Centers for Disease Control and Prevention (CDC) to assess youth behavior risk among high school students. Data collection procedures for the 2019 YRBS collection cycle were conducted under a continuation of the CDC institutional review boards protocol #1969.0. The sampling frame of the 2019 YRBS consisted of all public and non-public schools containing grades 9-12 and uses a three-stage cluster sample design to generate a nationally representative sample of high school students. For additional details on the YRBS sampling procedures refer to *Youth Risk Behavior Surveillance – United States, 2019* (CDC, 2018).

Items were selected based on relevance to the research question as indicated by prior literature and included (a) current age, (b) sexual experiences, including timing and age of initiation, number of sexual partners, current activity, history of sexual violence, and history of forced sexual contact, (c) substance use, including alcohol, nicotine, and illegal substances (d) symptoms of depression, (e) academic performance, (f) level of physical activity, and (g) race. Based on prior literature, a cutoff of at or below age 14 was utilized to determine early initiation of sexual intercourse (Cavazos-Rehg et al., 2009; Kaplan et al., 2013). Race was measured as White, Black or African American, Hispanic descent, all remaining races were combined into a single category to address small sample sizes among those who were American Indian, Alaskan Native, Asian, and Native Hawaiian or other Pacific Islander. To determine use of illegal substances, a dichotomous variable was created to indicate admission of use of any one of cocaine, ecstasy, heroin, inhalants, LSD, marijuana, methamphetamine, or synthetic marijuana. A dichotomous variable measuring having had multiple sex partners was created in order to differentiate between having had no more than one sexual partner versus having had multiple partners.

Data from all girls who responded to the 2019 National YRBS (*N* = 6,885) were analyzed using frequencies, crosstabulations, and logistic regression. All reported analyses were conducted by adjusting for the nature of the complex sampling process. As several states do not collect data on sexual behaviors, data could not be analyzed at the state level.

## Results

### Descriptive analyses for all girls

A complex samples crosstabulation between BMI and ESI using data from all girls (*N* = 6885) revealed similar findings for early initiation among girls who were classified as overweight (14.41% ± 1.68%) or obese (13.30% ± 1.48%) compared to those who were not (10.33% ± 0.69%). Further, among girls who reported having been physically forced to have sexual contact, a large percentage reported ESI (35.92% ± 2.78% vs 8.23% ± 0.57%), while a lower percentage were classified as a healthy weight (60.78% ± 2.97% vs 72.05% ± 1.54%).

A complex samples logistic regression revealed that girls who were overweight or obese were slightly more likely to initiate sex early compared to girls who were not (*OR* = 1.40, *95% CI:* 1.10, 1.78). And, when controlling for BMI classification, girls who had previously been physically forced to have sexual contact were 3.60 (*95% CI:* 2.42, 5.35) times more likely to initiate sex early compared to girls who had not experienced force. Given these findings, there appears to be a non-linear relationship between ESI and BMI that is modified by prior experience of forced sexual contact. To explore this relationship data were stratified by experiences of forced sexual contact for further analysis. Table 1 provides a descriptive representation of all girls who completed the 2019 YRBSS, as well as descriptive statistics comparing those who had (*n* = 630) or had not (*n* = 5009) reported prior experience of forced sexual contact.

**Table 1.**
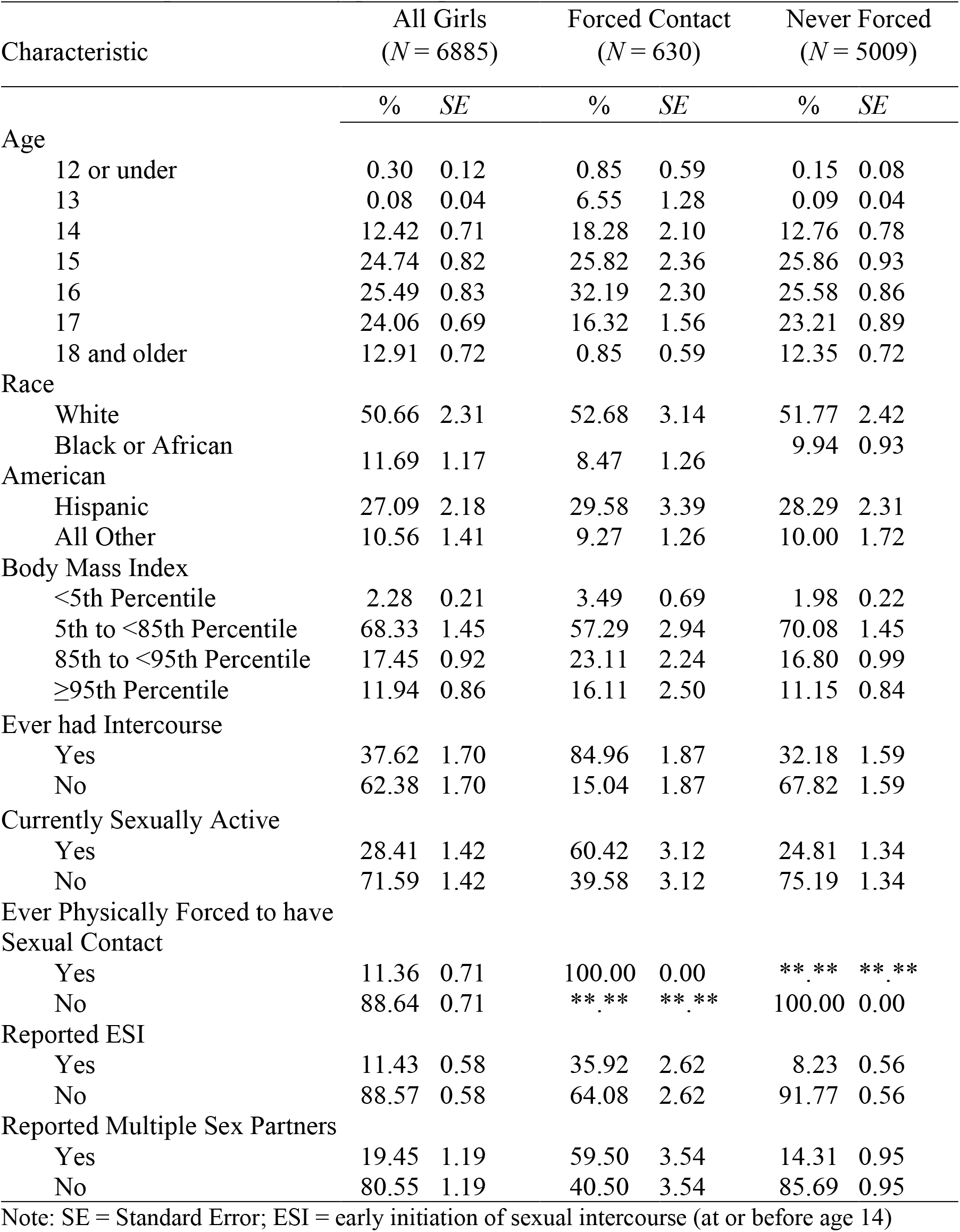
Descriptive statistics by prior experience with forced sexual intercourse.

### Girls who experienced forced sexual contact

Complex samples crosstabulation revealed a higher percentage of girls who were obese (44.38% ± 6.98%) initiated sex early compared to girls who were healthy weight (34.82% ± 3.82%) or overweight (31.77% ± 5.76%).

A complex samples logistic regression was conducted controlling for race, symptoms of depression, academic performance, illegal substance use, nicotine use, alcohol use, and level of physical activity. The analysis revealed that among girls who reported having experienced forced sexual contact those who were overweight (*OR* = 0.81, *95% CI:* 0.33, 2.03) or obese (*OR* = 1.49, *95% CI:* 0.62, 3.60) were no more likely to have initiated sex at or before age 14 than girls who were average or underweight. Similarly, there were no significant differences in early initiation found for ever having used illegal substances, current nicotine use, current alcohol consumption, symptoms of depression, or academic performance, see Table 2.

**Table 2.** Model summary comparing all girls to history of experiences of forced sexual intercourse.

| Variable | All girls |  | Forced Contact |  | Never Forced |  |
| --- | --- | --- | --- | --- | --- | --- |
|  | OR | 95% CI | OR | 95% CI | OR | 95% CI |
| **Race** |  |  |  |  |  |  |
| Black or African American | 1.68 | 1.06, 2.67 | 6.80* | 2.32, 19.96 | 1.19 | 0.58, 2.42 |
| Hispanic | 1.04 | 0.75, 1.43 | 0.96 | 0.41, 2.26 | 1.00 | 0.71, 1.40 |
| Other | 0.61 | 0.36, 1.02 | 0.68 | 0.23, 1.99 | 0.56* | 0.32, 0.99 |
| White (ref) | - | - | - | - | - | - |
| **BMI** |  |  |  |  |  |  |
| Overweight | 1.20 | 0.75, 1.94 | 0.81 | 0.33, 2.03 | 1.48 | 0.91, 2.40 |
| Obese | 1.10 | 0.73, 1.64 | 1.49 | 0.62, 3.60 | 0.87 | 0.45, 1.70 |
| Healthy Weight (ref) | - | - | - | - | - | - |
| **Symptoms of Depression** |  |  |  |  |  |  |
| Yes | 1.30 | 0.96, 1.78 | 2.22 | 0.89, 5.57 | 1.16 | 0.83, 1.62 |
| No (ref) | - | - | - | - | - | - |
| **Academic Performance** |  |  |  |  |  |  |
| Yes | 0.68 | 0.45, 1.03 | 0.92 | 0.41, 2.07 | 0.57* | 0.38, 0.88 |
| No (ref) | - | - | - | - | - | - |
| **Met Physical Activity Guidelines** |  |  |  |  |  |  |
| Yes | 1.02 | 0.73, 1.42 | 0.38* | 0.21, 0.69 | 1.30 | 0.86, 1.97 |
| No (ref) | - | - | - | - | - | - |
| **Current Nicotine Use** |  |  |  |  |  |  |
| Yes | 1.69* | 1.12, 2.55 | 1.34 | 0.62, 2.89 | 1.77* | 1.15, 2.72 |
| No (ref) | - | - | - | - | - | - |
| **Current Alcohol Use** |  |  |  |  |  |  |
| Yes | 1.14 | 0.82, 1.58 | 0.89 | 0.47, 1.70 | 1.32 | 0.92, 1.89 |
| No (ref) | - | - | - | - | - | - |
| **Prior Illicit Drug Use** |  |  |  |  |  |  |
| Yes | 2.38* | 1.61, 3.54 | 1.15 | 0.59, 2.23 | 2.89* | 1.77, 4.73 |
| No (ref) | - | - | - | - | - | - |
| **Forced Sexual Contact** |  |  |  |  |  |  |
| Yes | 3.60* | 2.42, 5.35 | - | - | - | - |
| No (ref) | - | - | - | - | - | - |

Notes: \* = Statistically significant; ref = reference group; Symptoms of Depression = reported feeling sad or hopeless over a two-week period; Academic performance = described grades as mostly A's and B's;

However, girls who were ‘Black or African American’ and reported having experienced forced contact were 6.80 (2.31, 19.96) times more likely to report ESI than girls who were ‘White’. While girls who were engaging in physical activity were less likely to report ESI than girls who were not (*OR* = 0.38, *95% CI:* 0.21, 0.69), as well as for girls who reported current use of nicotine (*OR* = 1.77, *95% CI:* 1.15, 2.72). All other comparisons can be seen in Table 2.

### Girls who never experience forced sexual contact

Complex samples crosstabulation revealed a similar percentage of girls who were healthy weight (7.64% ± 0.69%) and obese (6.61% ± 1.69%) initiated sex early compared to 11.55% (1.70) of girls who were overweight.

A complex samples logistic regression was conducted controlling for race, symptoms of depression, academic performance, illegal substance use, nicotine use, alcohol use, and level of physical activity. The analysis revealed that among girls who reported having never experienced forced sexual contact those who were overweight (*OR* = 1.48, *95% CI:* 0.91, 2.40) or obese (*OR* = 0.87, *95% CI:* 0.45, 1.70) were no more likely to have initiated sex at or before age 14 than girls who were average or underweight. Similarly, there were no significant differences in early initiation found for symptoms of depression, current alcohol consumption, or physical activity, see Table 2.

However, girls who reported having ever used an illegal substance were more likely to report ESI than girls who had not (*OR* = *2*.89, *95% CI:* 1.77, 4.73), as well as for girls who reported current use of nicotine (*OR* = 1.77, *95% CI:* 1.15, 2.72). While girls who reported earning grades of “As and Bs” were 0.57 (0.38, 0.88) times less likely to report ESI than girls who earned lower grades and those who reported a race or ethnicity other than ‘Black or African American’ or ‘Hispanic’ were 0.56 (0.32, 0.99) times less likely to report ESI than girls who were ‘White’. All other comparisons can be seen in Table 2.

## Discussion

The current study differs from prior research in that stratification was used to isolate the impact of sexual trauma, a known predictor of increased BMI (Davis et al., 2019; Opel et al., 2019) and early sexual initiation (Greydanus et al., 2010; McCutcheon et al., 2017). The main results of this study indicate that girls with a weight status of overweight or obese were no more likely to initiate sex early compared to girls who were average or underweight when forced sexual contact was included in the analysis. This indicates that weight status may not be a contributing factor to ESI for girls. Thus, these results do not support BMI as one of the myriads of variables that interact to influence sexual health related behaviors, but rather that having experienced an adverse childhood event (ACE) likely contributed to poor health choices.

Adolescent girls who had experience forced sexual intercourse were much more likely to initiate early sexual contact than girls who had not and controlling for this effect altered the nature of the relationship between other predictors and ESI. When girls had never experienced force, those who reported current nicotine use or who reported having ever used an illegal substance were more likely to also report ESI, indicating that early exposure to substance use may be a leverage point for intervention to prevent ESI among these girls. However, when girls reported having experienced force, girls who were Black or African American where especially at risk of reporting ESI despite a similar distribution of race in both groups. The effect of illicit drug and nicotine use vanished when controlling for experience of force, but this was likely due to a large proportion of the adolescent girls in this category reporting experience with substance use.

There is a myriad of factors that contribute to ESI and these factors may differ for girls who have or have not experienced forced sexual intercourse. The results of this study support the findings of McCutcheon et al. (2017) and Davis et al. (2019) indicating that ACEs are related to increases in BMI as well ESI. Prior literature also supports that the more ACEs a girl experiences the more likely she is to engage in ESI (Hillis et al., 2001). Ultimately, while BMI and ESI may occur together, BMI does not impact ESI. As indicated by Greydanus et al. (2010), forced intercourse has a known effect on ESI, and appears to be a primary predictor of early initiation as well as other risky sexual behaviors such as having multiple partners.

### Study limitations

One of the main limitations, is the lack of consistency in the way in which early sexual initiation or sexual debut is defined (National Center for Health Statistics, 2017; Kaplan et al., 2013; CDC, 2018). The term denotes initiating sex at a young age, but the extent to which that age is defined varies and can lead to a lack of consistency when comparing research in this area. The YRBS is cross-sectional in nature making it impossible to determine directionality. Therefore, it cannot be established if forced sexual contact occurred prior to (or post) sexual initiation. It is also important to note that the questions of the YRBS do not instruct respondents to disregard sexual assault or abuse when determining the age at which they initiated sex. This may very well be the cause of the lack of association found between weight status and ESI among girls who experienced forced sex.

Another limitation is the extent to which adolescents understand the term “sexual intercourse”. The YRBS does not provide a definition of sexual intercourse. Instead, it is assumed that adolescents understand this terminology. Some participants in the YRBS may be 12 years old and younger and have limited knowledge and understanding of the term “sexual intercourse”. While we may assume that older adolescents, such as those near the end range of age in the sample, understand what sexual intercourse it is also safe to assume that some don’t.

When the lack of comprehensive sexual education is accounted for in many areas of the country, the extent to which “sexual intercourse” is clearly understood can be called into question further (Foley, 2015).

### Implications for practice

The results of this study can better assist public health professionals by providing them with the knowledge that BMI is not predictive of ESI among girls. However, the ACE of forced sexual intercourse is strongly predictive of ESI and leverage points for intervention may provide opportunities the mitigate the deleterious effect of this ACE. This study can also aide in reducing the stigmatizing belief that obese adolescent girls are more likely to participate in ESI.

## Data Availability

All data produced are available online at: https://www.cdc.gov/healthyyouth/data/yrbs/index.htm

